# Motivational Strategies for Stroke Rehabilitation: A Delphi Study

**DOI:** 10.1101/2020.03.15.20036764

**Authors:** Kazuaki Oyake, Makoto Suzuki, Yokei Otaka, Kimito Momose, Satoshi Tanaka

## Abstract

**Background and Purpose:** Although various strategies are used to motivate patients during rehabilitation, consensus regarding the optimal motivational strategies for stroke rehabilitation has not been established. Expert consensus may aid rehabilitation professionals in effectively utilizing motivational strategies to produce the most beneficial outcome for their patients. The primary purpose of this study was to provide a comprehensive list of effective motivational strategies based on consensus among rehabilitation experts, generated using the Delphi technique. In addition, we sought to identify the types of information that are important when selecting motivational strategies.

**Methods:** A total of 198 rehabilitation experts participated in a three-round Delphi survey. The rehabilitation experts included physicians, physical therapists, occupational therapists, and speech-language-hearing therapists who had worked in stroke rehabilitation for at least five years. Panelists were asked to rate the effectiveness of motivational strategies and to rate the importance of different types of information using a 5-point Likert scale. Consensus was defined as having been reached for items with an interquartile range of 1 or less.

**Results:** A total of 116 experts (58.6%) completed the third round of the Delphi survey. Consensus was reached on all of the 26 presented strategies. Seven strategies, such as control of task difficulty and goal setting, were considered to be very effective in increasing patient motivation. In addition, all 11 of the presented types of information were deemed very important or important in determining which motivational strategies to use.

**Conclusions:** We generated a list of effective motivational strategies for stroke rehabilitation based on expert consensus. Our results suggest that experts consider a comprehensive range of patient information when choosing motivational strategies. These findings represent a group of consensus-based recommendations for increasing patient adherence to stroke rehabilitation programs, which may be beneficial to many medical professionals working in stroke rehabilitation.

## Introduction

Rehabilitation programs, such as a repetitive task-specific practice and an exercise training, can improve functional capacity, the ability to perform activities of daily living, and quality of life.^1^ High adherence to a prescribed rehabilitation program is regarded as indicative of motivation.^2^ Rapolienė et al. (2018)^3^ found that higher internal motivation at the beginning of a rehabilitation program was correlated with the greater improvement in the activities of daily living in stroke patients. Therefore, the addition of motivational strategies to rehabilitation programs may promote functional improvement after stroke.^4^

Various strategies are used to motivate stroke patients in clinical practice.^2, 5^ One study using a semi-structured interview format reported that rehabilitation professionals set rehabilitation goals, provided information regarding rehabilitation, and accessed and used the patient’s cultural norms to increase patient motivation.^2^ In addition, we previously found that 15 motivational strategies, such as active listening and praise, were used by more than 75% of 362 professionals working in stroke rehabilitation.^5^ Indeed, motivational strategies can positively affect rehabilitation in stroke patients.^6-8^ For example, an international randomized clinical trial indicated that encouragement effectively improved walking speed in stroke patients.^6^ However, there is no published consensus regarding the most effective motivational strategies or the importance of specific types of information during the selection of motivational strategies. Expert consensus about which strategies are considered effective and how they are selected will help rehabilitation professionals to increase patient adherence to stroke rehabilitation programs and improve patient outcome. The Delphi method is a well-known approach for developing expert consensus regarding topics where published evidence is scare.^9^

The purpose of this study was to use the Delphi method to generate a list of effective motivational strategies based on consensus among rehabilitation experts. We hypothesized that rehabilitation experts may use different motivational strategies according to the patient’s condition. Therefore, we also aimed to identify the types of information used when selecting motivational strategies.

## Methods

### Data availability statement

The data that support the findings of this study are available from the corresponding author upon reasonable request.

### Study design

We conducted a Delphi study according to the guidance on conducting and reporting Delphi studies (CREDES).^10^ Three iterations are typically though to be sufficient to identify points of consensus.^10-12^ Therefore, the total number of Delphi rounds was set at three.

### Participants

The Delphi method is an anonymous iterative survey of exert opinion.^13^ Thus, eligible participants, recruited via a convenience sampling method, were experts in stroke rehabilitation. In this study, experts were defined as physicians, physical therapists, occupational therapists, and speech-language-hearing therapists who had worked in stroke rehabilitation for at least five years. Experts were recruited with the cooperation of the 17th Annual Academic Conference of the Japanese Society of Neurological Physical Therapy and the 29th Annual Meeting of the Japan Society for Respiratory Care and Rehabilitation. We set up a booth inside each conference and provided laptops that conference goers could use to voluntarily access the website that contained the questionnaire (the Google Forms tool, Google LLC, Mountain View, CA, USA). Potential participants could also access the survey website using their laptops, tablets, or smartphones from a hyperlink given in a leaflet and displayed on a poster in front of the booth. In addition, we sent email invitations to experts who participated in our previous study.^5^ On the first page of the website, participants were asked to report their professional characteristics, such as professional category and years of experience working in rehabilitation. Those who met the eligibility criteria could participate in the Delphi survey. According to the Consensus-based Standards for the selection of health Measurement Instruments (COSMIN) recommendations,^14^ we aimed to recruit a total of at least 120 experts in the first round of the survey.

### Ethical considerations

The study was approved by the appropriate ethics committees at the Hamamatsu University School of Medicine (approval number: 19-048) and at the Shinshu University (approval number: 4385). Informed consent was obtained from all participants.

### Developing the list of motivational strategies

In this study, we defined motivational strategies as concrete tactics, techniques, or approaches to orient patients to rehabilitation.^15, 16^ We expected this definition to produce a large list of items used by rehabilitation professionals, while some of the presented strategies might be overlapped with one another. Rather than strictly define motivational strategies, we emphasized the applicability of the strategies in a clinical setting. Although some of the included motivational strategies might be difficult to distinguish from elements of good clinical practice (e.g., active listening and praise), they were included because they have been considered to enhance patient participation in rehabilitation.

We initially developed a list of motivational strategies based on data obtained from semi-structured interviews,^17^ the findings of related literature,^2, 6, 18-24^ and the clinical experience of the authors. First, we obtained a list of 10 motivational strategies, such as goal setting and praise, from the semi-structured interviews.^17^ Then, we identified six strategies, including providing medical information and having family members present during rehabilitation, from the related previous studies.^2, 6, 18-24^ We then added six strategies, such as active listening and goal-oriented practice, to the list based on our clinical experience. The content validity of the items was assessed through expert review and a pilot test,^25^ which resulted in some minor grammatical changes. Using the resulting list of 22 motivational strategies, we surveyed more than 360 rehabilitation professionals regarding the motivational strategies commonly used in clinical practice for stroke rehabilitation.^5^ The survey included an open-ended question in which respondents were invited to propose additional motivational strategies not included on the list. As a result of the survey, we added two motivational strategies (“providing feedback to the patient regarding the results of the practice” and “engage in practice with the patient”) to the list. Finally, we also added motivational interviewing and cognitive behavioral therapy to the list because these strategies were regarded as the package of motivational interventions in the previous studies.^4, 26^ Consequently, we prepared a list of 26 motivational strategies (Table 1).

**Table 1.**
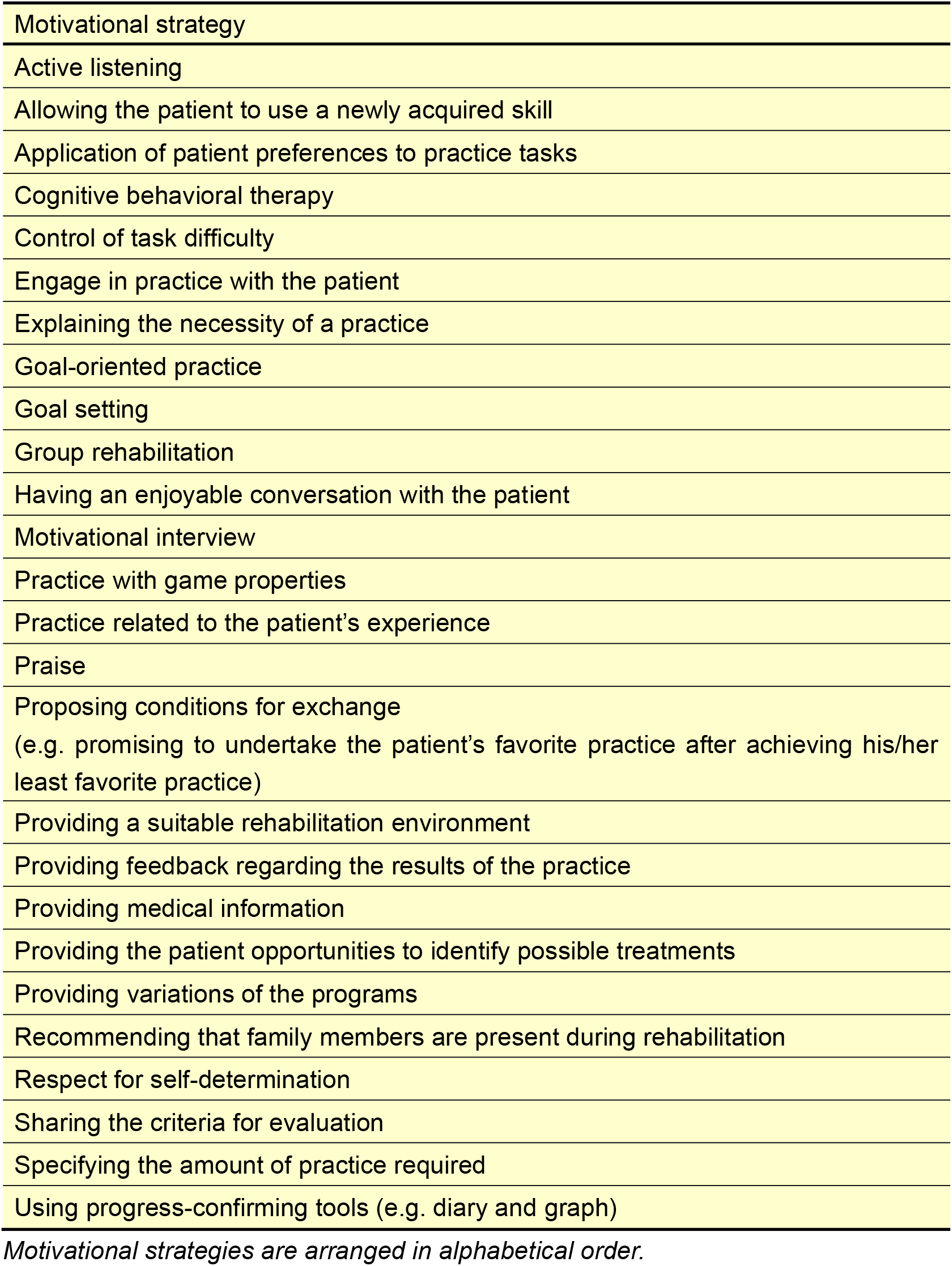
List of motivational strategies.

In addition, we prepared a list of 11 type of information regarding the patient’s health status, environmental factors, and personal factors (Table 2). We used this list to identify which types of information are considered to be important when selecting motivational strategies.

**Table 2.** List of types of information considered by rehabilitation professionals when choosing motivational strategies.

|  |
| --- |
| Type of information |
| Cognitive function (e.g. higher brain dysfunction and dementia) |
| Comorbidities<br>(e.g. psychological disorders, diabetes mellitus, and infection) |
| Demographic characteristics (e.g. age and sex) |
| Diagnosis (e.g. ischemic or hemorrhagic, lesion site, and recurrence) |
| Human environment (e.g. key person and family) |
| The patient's reaction to a presented motivational strategy |
| Personality |
| Physical function<br>(e.g. muscle weakness and limited range of motion) |
| The severity of activity limitations |
| The severity of participation restrictions |
| Social environment<br>(e.g. economic condition and employment status) |

*Information types are arranged in alphabetical order.*

### The Delphi method

The Delphi survey was conducted between September and December 2019. The first round survey was conducted between September and November 2019, and the second round questionnaire was sent in November 2019. The third round questionnaire was sent in December 2019. The second and third round questionnaires were sent to panelists who completed the questionnaire from the previous round. To maximize response rates, reminder emails were sent one and two weeks after each questionnaire was sent. In addition, panelists who completed the third round questionnaire received 1,000 yen (approximately US $9.00) honorarium.

In the first round, panelists were presented with 25 of the motivational strategies and the 11 types of patient information. “Active listening” was not included in the list of motivational strategies in the first round due to a technical error. It was added to the list in the second and third rounds. Panelists were asked to rate the effectiveness of each motivational strategy and the importance of each type of information using a 5-point Likert scale (where 1 = very ineffective/not at all important, 2 = ineffective/unimportant, 3 = uncertain, 4 = effective/important, and 5 = very effective/very important).

In the second and third rounds, the lists of 26 motivational strategies (including “active listening”) and the 11 types of information were again presented to the panelists. The response data for each strategy and type of information from the previous round were added as a reference, including a pie chart showing the distribution of responses and the mean rating score. All panelists were asked to reconsider their previous ratings and to rate each item again.

### Data analysis

To examine the effectiveness of each motivational strategy and the importance of each type of information, we calculated the median and interquartile range on a 5-point Likert scale for each item. An interquartile range of less than 1 indicates that more than 50% of all responses fall within 1 point on the scale.^27^ Items with an interquartile range of 1 or less are considered to demonstrate good consensus on a 5-point Likert scale.^12, 28^ Therefore, consensus was defined as an interquartile range of 1 or less.

## Results

### Participants

Of the 198 rehabilitation experts who accessed the survey website, 160 experts (80.8%) completed the first round of the survey. The panelists characteristics are shown in Table 3. The majority of participants were physical therapists (76.9%). Responses were obtained from 123 (62.1%) and 116 (58.6%) panelists in the second and third rounds, respectively. The flow of participants during the three rounds of the Delphi survey is shown in Figure 1.

**Table 3.**
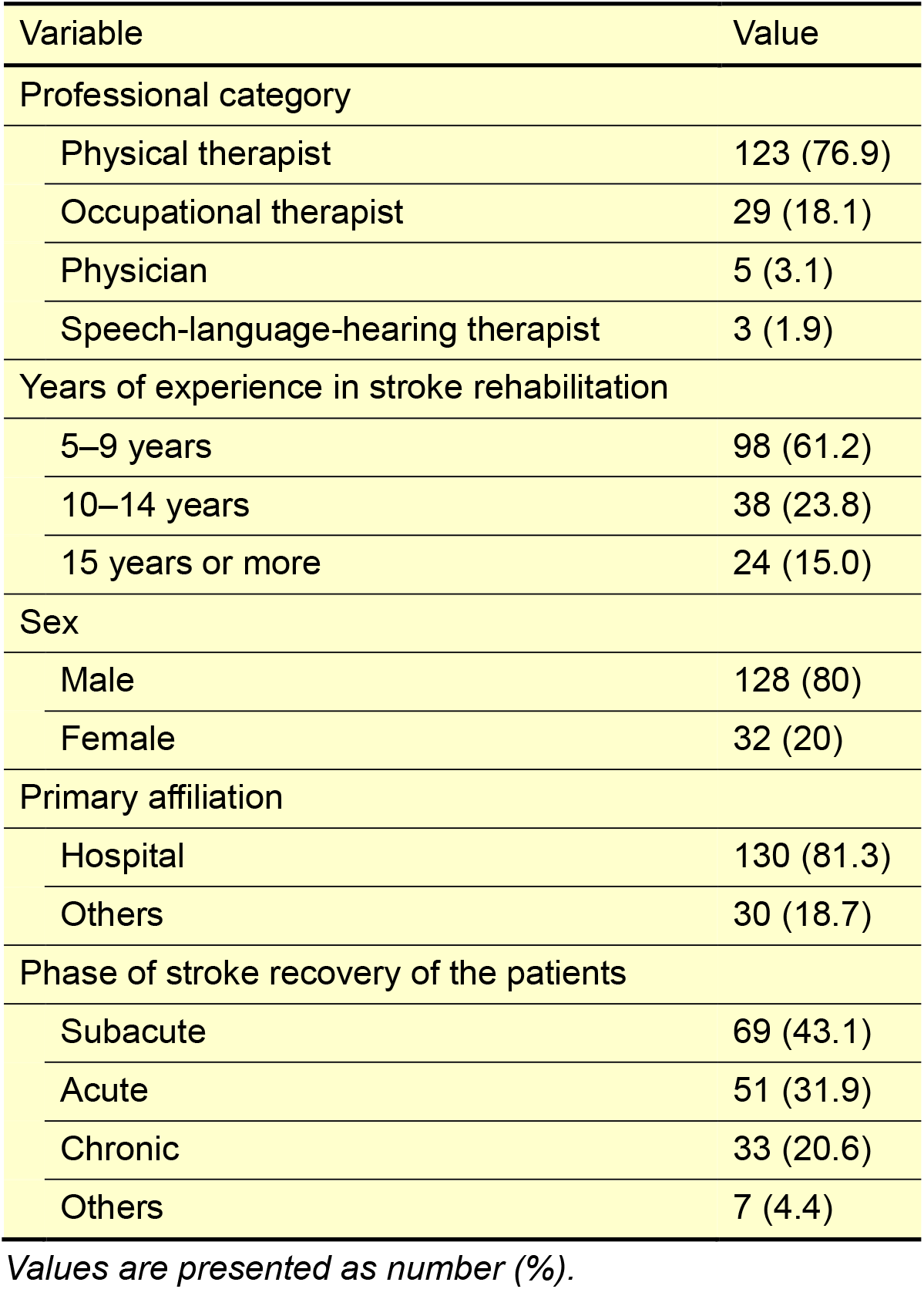
Characteristics of panelists (n = 160)

| Variable | Value |
| --- | --- |
| Professional category |  |
| Physical therapist | 123 (76.9) |
| Occupational therapist | 29 (18.1) |
| Physician | 5 (3.1) |
| Speech-language-hearing therapist | 3 (1.9) |
| Years of experience in stroke rehabilitation |  |
| 5–9 years | 98 (61.2) |
| 10–14 years | 38 (23.8) |
| 15 years or more | 24 (15.0) |
| Sex |  |
| Male | 128 (80) |
| Female | 32 (20) |
| Primary affiliation |  |
| Hospital | 130 (81.3) |
| Others | 30 (18.7) |
| Phase of stroke recovery of the patients |  |
| Subacute | 69 (43.1) |
| Acute | 51 (31.9) |
| Chronic | 33 (20.6) |
| Others | 7 (4.4) |

*Values are presented as number (%).*

**Figure 1.**
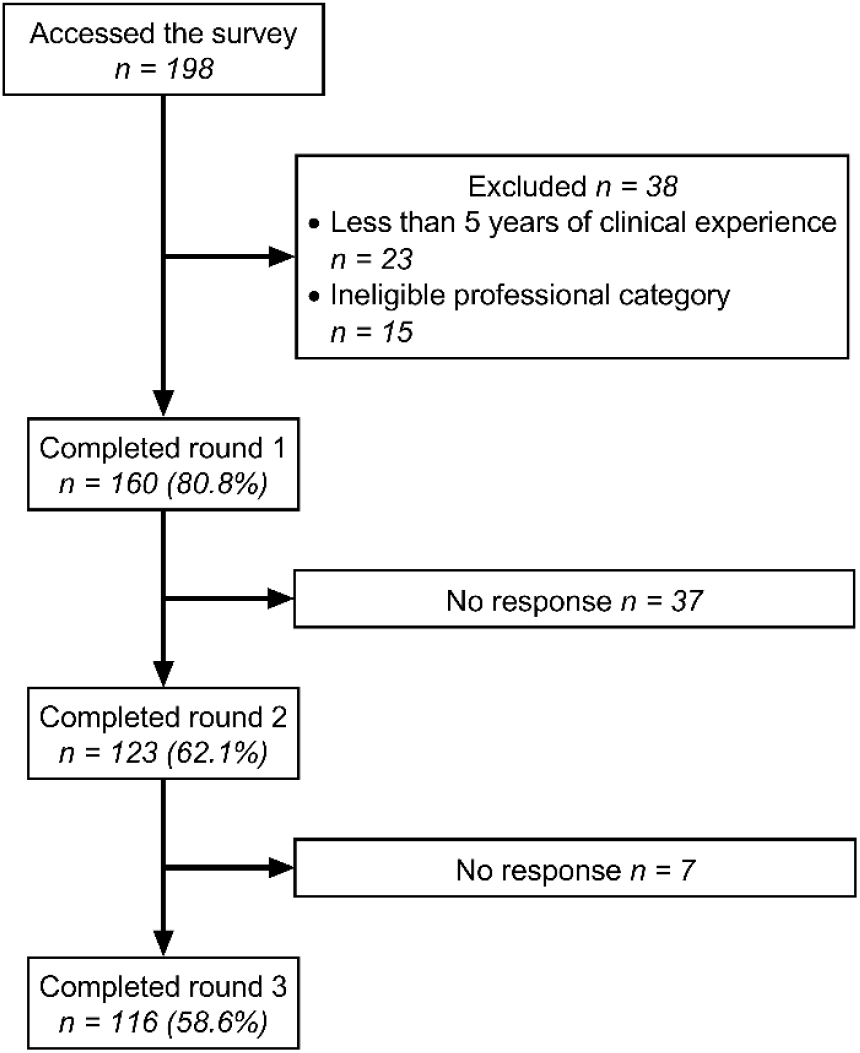
Flow diagram of the panelist response rates and withdrawals by round.

### Which motivational strategies did experts deem effective for stroke rehabilitation

The results of the Delphi survey on the effectiveness of each motivational strategy for stroke rehabilitation are shown in Table 4 and Supplemental Figure I. In the first round, consensus was achieved regarding 22 of the strategies. “Control of task difficulty”, “goal setting”, and “providing feedback to the patient regarding the results of the practice” were deemed very effective motivational strategies. Nineteen strategies, including “goal-oriented practice” and “praise”, were considered effective in motivating patients with stroke to engage in rehabilitation practices.

**Table 4.**
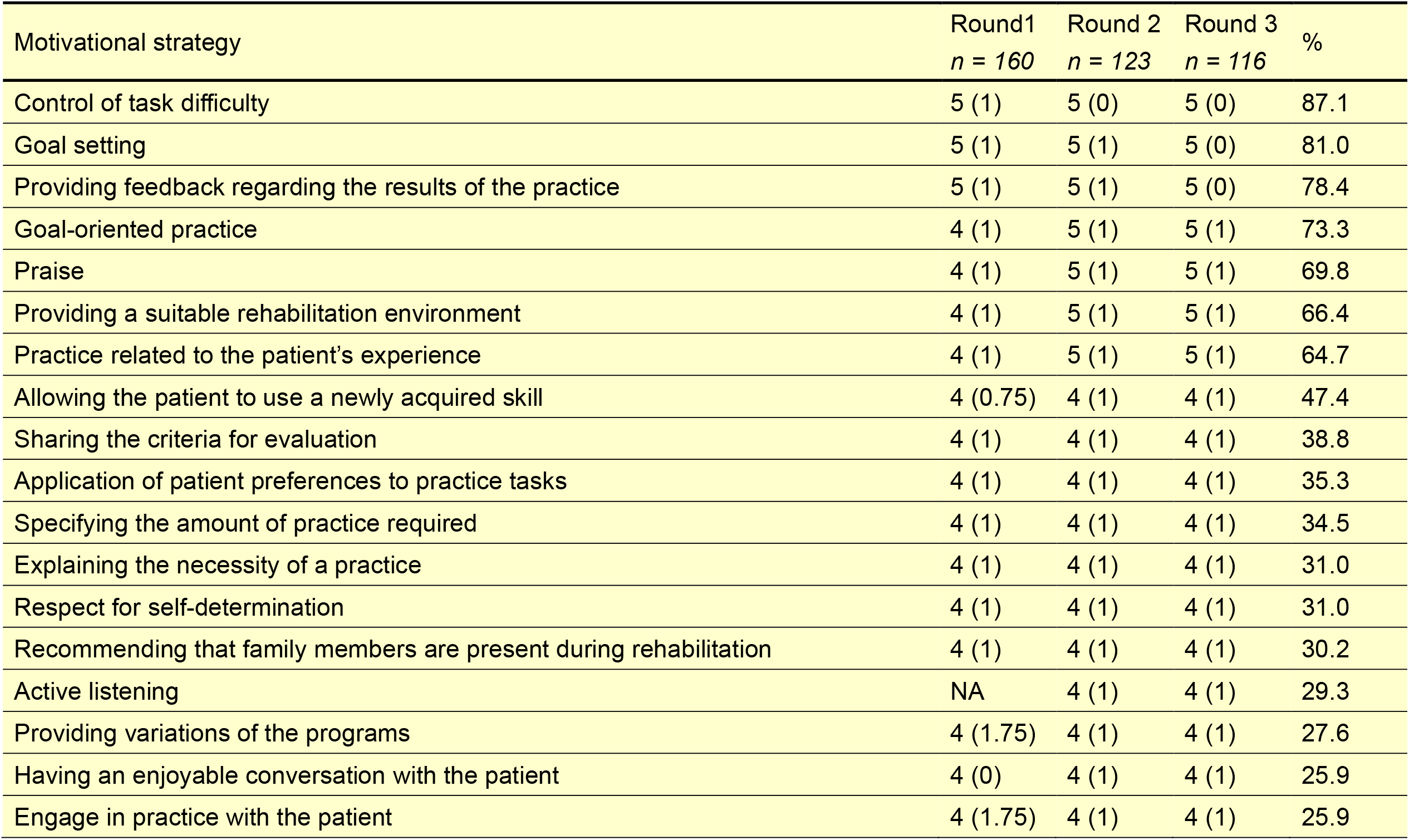
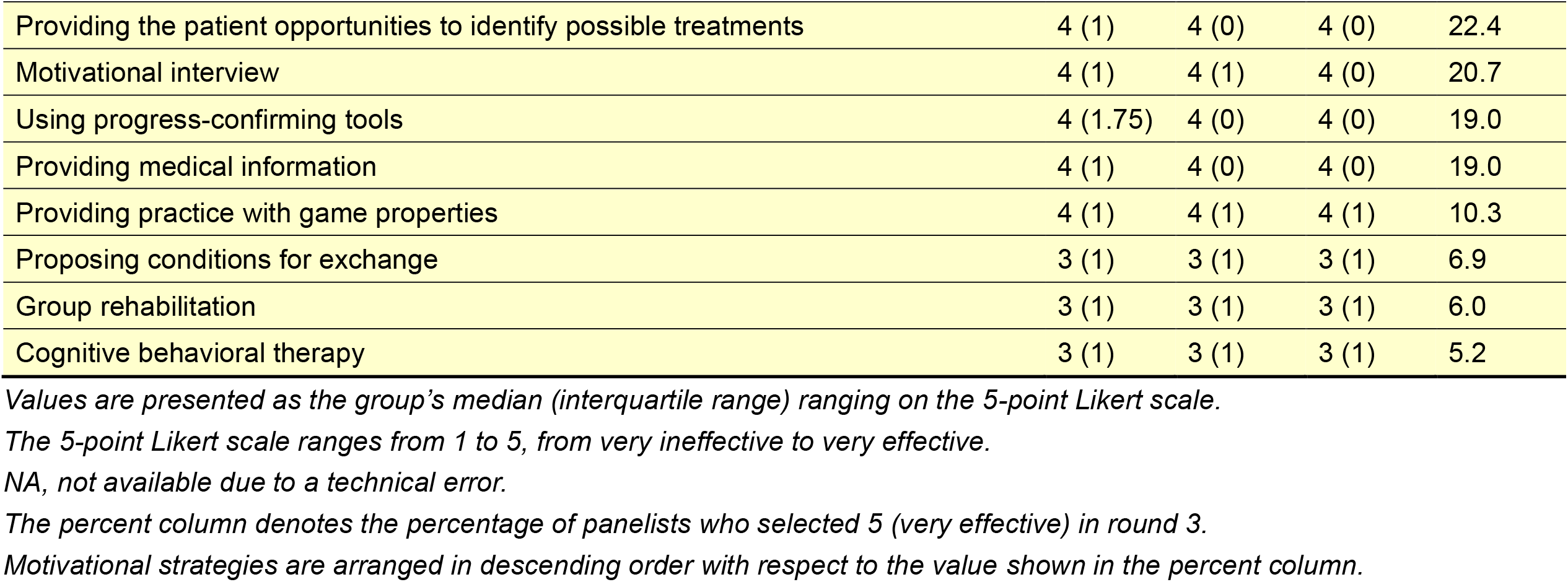
The effectiveness of each motivational strategy for stroke rehabilitation.

| Motivational strategy | Round1<br>*n* = 160 | Round 2<br>*n* = 123 | Round 3<br>*n* = 116 | % |
| --- | --- | --- | --- | --- |
| Control of task difficulty | 5 (1) | 5 (0) | 5 (0) | 87.1 |
| Goal setting | 5 (1) | 5 (1) | 5 (0) | 81.0 |
| Providing feedback regarding the results of the practice | 5 (1) | 5 (1) | 5 (0) | 78.4 |
| Goal-oriented practice | 4 (1) | 5 (1) | 5 (1) | 73.3 |
| Praise | 4 (1) | 5 (1) | 5 (1) | 69.8 |
| Providing a suitable rehabilitation environment | 4 (1) | 5 (1) | 5 (1) | 66.4 |
| Practice related to the patient's experience | 4 (1) | 5 (1) | 5 (1) | 64.7 |
| Allowing the patient to use a newly acquired skill | 4 (0.75) | 4 (1) | 4 (1) | 47.4 |
| Sharing the criteria for evaluation | 4 (1) | 4 (1) | 4 (1) | 38.8 |
| Application of patient preferences to practice tasks | 4 (1) | 4 (1) | 4 (1) | 35.3 |
| Specifying the amount of practice required | 4 (1) | 4 (1) | 4 (1) | 34.5 |
| Explaining the necessity of a practice | 4 (1) | 4 (1) | 4 (1) | 31.0 |
| Respect for self-determination | 4 (1) | 4 (1) | 4 (1) | 31.0 |
| Recommending that family members are present during rehabilitation | 4 (1) | 4 (1) | 4 (1) | 30.2 |
| Active listening | NA | 4 (1) | 4 (1) | 29.3 |
| Providing variations of the programs | 4 (1.75) | 4 (1) | 4 (1) | 27.6 |
| Having an enjoyable conversation with the patient | 4 (0) | 4 (1) | 4 (1) | 25.9 |
| Engage in practice with the patient | 4 (1.75) | 4 (1) | 4 (1) | 25.9 |
| Providing the patient opportunities to identify possible treatments | 4 (1) | 4 (0) | 4 (0) | 22.4 |
| Motivational interview | 4 (1) | 4 (1) | 4 (0) | 20.7 |
| Using progress-confirming tools | 4 (1.75) | 4 (0) | 4 (0) | 19.0 |
| Providing medical information | 4 (1) | 4 (0) | 4 (0) | 19.0 |
| Providing practice with game properties | 4 (1) | 4 (1) | 4 (1) | 10.3 |
| Proposing conditions for exchange | 3 (1) | 3 (1) | 3 (1) | 6.9 |
| Group rehabilitation | 3 (1) | 3 (1) | 3 (1) | 6.0 |
| Cognitive behavioral therapy | 3 (1) | 3 (1) | 3 (1) | 5.2 |

*Values are presented as the group's median (interquartile range) ranging on the 5-point Likert scale.*
*The 5-point Likert scale ranges from 1 to 5, from very ineffective to very effective.*
*NA, not available due to a technical error.*
*The percent column denotes the percentage of panelists who selected 5 (very effective) in round 3.*
*Motivational strategies are arranged in descending order with respect to the value shown in the percent column.*

In the second round, consensus was reached for all of the 26 presented strategies. In addition to the three strategies considered very effective in the first round, “goal-oriented practice”, “praise”, “providing a suitable rehabilitation environment”, and “practice related to the patient’s experience” were also regarded as very effective motivational strategies. Sixteen strategies were considered effective motivational strategies. The results of the third round were similar to those obtained in the second round. “Proposing conditions for exchange”, “group rehabilitation”, and “cognitive behavioral therapy” were considered neither effective nor ineffective during the three rounds.

### Which types of information do experts consider important when they choose motivational strategies

The results of the Delphi survey on the importance of different types of information when selecting motivational strategies are shown in Table 5 and Supplemental Figure II. In the first round, consensus was reached for 9 of the presented 11 items. “Cognitive function” and “personality” were considered to be very important types of information. Seven types of information were deemed important.

**Table 5.** The importance of each type of information when selecting motivational strategies.

| Type of information | Round1<br>*n* = 160 | Round 2<br>*n* = 123 | Round 3<br>*n* = 116 | % |
| --- | --- | --- | --- | --- |
| Cognitive function | 5 (1) | 5 (0) | 5 (0) | 85.3 |
| Human environment | 4 (1) | 5 (1) | 5 (1) | 66.3 |
| Personality | 5 (1) | 5 (1) | 5 (1) | 59.5 |
| The patient's reaction to a presented motivational strategy | 4 (1) | 5 (1) | 5 (1) | 56.9 |
| Social environment | 4 (1) | 4 (1) | 5 (1) | 54.3 |
| The severity of activity limitations | 4 (1) | 4 (1) | 5 (1) | 50.9 |
| Physical function | 4 (1) | 4 (1) | 4 (1) | 44.0 |
| Comorbidities | 4 (1) | 4 (1) | 4 (1) | 41.4 |
| Diagnosis | 4 (2) | 4 (1) | 4 (1) | 35.3 |
| The severity of participation restrictions | 4 (1) | 4 (1) | 4 (1) | 25.9 |
| Demographic characteristics | 4 (2) | 4 (1) | 4 (0) | 21.6 |

*Values are presented as the group's median (interquartile range) ranging on the 5-point Likert scale.*
*The 5-point Likert scale ranges from 1 to 5, from not at all important to very important.*
*NA, not available due to a technical error.*
*The percent column denotes the percentage of panelists who selected 5 (very important) in round 3.*
*Types of information are arranged in descending order with respect to the value shown in the percent column.*

In the second round, consensus was achieved for all 11 types of information. “Human environment” and “the patient’s reaction to a presented motivational strategy”, “cognitive function”, and “personality” were considered very important. The other seven types of information were regarded as important.

After the third round, “social environment” and “the severity of activity limitations” were also deemed very important, while five other types of information were considered important.

## Discussion

We generated a list of motivational strategies for stroke rehabilitation based on expert consensus. We identified 23 strategies that were deemed very effective or effective in motivating patients with stroke to engage in rehabilitation practices. Our results show that rehabilitation experts consider a comprehensive range of information regarding the patient’s health condition, environmental factors, and personal factors when choosing motivational strategies. These findings can be used to generate consensus-based recommendations regarding motivational strategies for stroke rehabilitation.

To the best of our knowledge, this study is the first to establish consensus among experts regarding motivational strategies for stroke rehabilitation using the Delphi method. Although expert opinion is regarded as the lowest grade of evidence,^29^ the Delphi process can be used to enhance the authority of expert opinion.^30^ In addition, the Delphi method can be used to generate an evidence base in situations where evidence is insufficient.^9^ As evidence-based motivational strategies have not yet been established with respect to stroke rehabilitation, a consensus of expert opinion generated using the Delphi method may be beneficial in helping rehabilitation professionals to enhance patient motivation. According to the COSMIN recommendations, a rating of ‘very good’ can be established if the sample size was at least 100.14 Diamond et al. (2014)^11^ reported that only 5 of 100 studies using the Delphi method recruited more than 100 panelists in the final round. Therefore, that we obtained data from 116 experts in the final round is an important strength of this study.

### Which motivational strategies did experts deem effective for stroke rehabilitation

We found seven very effective and 16 effective strategies for motivating patients with stroke to engage in rehabilitation practices. Of the seven very effective strategies, all except for ‘providing feedback to the patient regarding the results of the practice’ were used by more than 75% of the rehabilitation professionals in our previous study.^5^ Gradual increases in task difficulty and goal-oriented practice have been recommended to promote functional recovery during stroke rehabilitation.^31^ As inappropriate levels of task difficulty can bore or frustrate the patient,^32^ the difficulty of a practice task is considered to affect the patient adherence to a rehabilitation program. Setting rehabilitation goals can also have a positive impact on stroke recovery during rehabilitation.^33^ In addition, positive feedback and encouragement have been reported to be effective for improving walking speed during inpatient rehabilitation.^6^ Sonoda et al. (2004)^34^ found that a specific rehabilitation environment helped stroke inpatients increase their amount of physical activity during daily living. Therefore, these strategies appear to be very effective for motivating patients with stroke during rehabilitation.

Self-determination facilitates and maintains intrinsic motivation.^35^ Active listening is a core communication skill involved in motivational interviewing,^7, 26^ and undergoing motivational interviewing has been associated with normal mood.^7^ The provision of information regarding rehabilitation and the presence of a family member during rehabilitation may improve a patient’s mood and encourage them to be more active.^8, 36^ Communication training programs for clinicians increase patient satisfaction, levels of motivation for goal setting and action, and health-related quality of life.^37^ Rehabilitation programs with game properties such as virtual reality have been found to improve motivation and activities of daily living.^38, 39^ Thus, some strategies considered to be effective have been shown to have positive effects on recovery after stroke.

### Which types of information do experts consider important when they choose the motivational strategies

The findings of this study support previous findings regarding factors related to motivation in stroke patients.^5, 21, 22, 24, 40^ Our results suggest that the following are regarded as particularly essential information when choosing motivational strategies: the severity of cognitive impairments and activity limitations, human and social environments, patient personality type, and patient responses to motivational strategies. For example, in the authors’ clinical experience, providing a patient with severe cognitive impairments and/or activity limitations with a relatively easy practice task can help to maintain patient confidence. In addition, rehabilitation professionals must consider the social environment and personality of a patient when selecting tasks for inclusion in a goal-oriented program, and design the practice with consideration of patient preferences. Furthermore, our results suggest that experts adjust their motivational strategies according to patient reactions. In the present study, we did not classify the strategies according to patient condition, such as neurological status or personality. Therefore, further studies are needed to evaluate the motivational strategies that rehabilitation experts use according to patient condition.

### Limitations

There exist several limitations to the present study. First, all of the panelists were recruited in Japan. Thus, whether our findings are generalizable to rehabilitation professionals outside of Japan remains unclear. For instance, cognitive behavioral therapy was rated as neither effective nor ineffective by the experts in the present study. Our supplemental survey showed that less than 50% of the panelists (45.6%) were familiar with cognitive behavioral therapy. Thus, the low ratings of effectiveness for cognitive behavioral therapy were likely due to a low degree of recognition among rehabilitation experts in Japan. With the exception of our data for cognitive behavioral therapy, our results are consistent with those of some previous qualitative^2, 21, 22, 24, 40^ and experimental^6-8, 33, 34, 36-39^ studies from Western countries. An international Delphi survey would improve the external validity of our findings.

Second, we utilized convenience sampling to recruit the panelists in this study. We recruited individuals for voluntary participation by distributing leaflets, displaying posters, and sending email invitations. Thus, the experts who participated in this study may have been more interested in the motivational strategies compared with nonparticipants. Therefore, the effectiveness of each motivational strategy may be overestimated, and so caution is warranted in interpreting our results. Third, the majority of the participants were physical therapists. As a result, the opinions of physical therapists might be overstated in the data. There are approximately 172 thousand (as of March 2019),^41^ 80 thousand (as of March 2017),^42^ 30 thousand (as of March 2019),^43^ and 2 thousand (as of June 2017)^44^ licensed physical therapists, occupational therapists, speech-language-hearing therapists, and rehabilitation doctors in Japan, respectively. Thus, the large population of physical therapists in our sample appears to be consistent with the actual situation in Japan. Nonetheless, in future research, a random sampling method might be useful in order to minimize this type of sampling bias.

## Summary

To the best of our knowledge, this is the first study to generate a list of effective motivational strategies for stroke rehabilitation based on expert consensus. Seven motivational strategies, including controlling task difficulty and setting rehabilitation goals were deemed very effective. In addition, 16 strategies, such as allowing the patient to use a newly acquired skill and sharing the criteria for evaluation, were considered effective in increasing patient motivation. Our results suggest that experts consider a comprehensive range of data regarding patient condition when making decisions about motivational strategies for rehabilitation. These findings may be useful for developing consensus-based recommendations regarding motivational strategies for use in stroke rehabilitation.

## Acknowledgments

The authors thank Sydney Koke, MFA, from Edanz Group (www.edanzediting.com/ac) for editing a draft of this manuscript.

## References

1. Billinger SA, Arena R, Bernhardt J, Eng JJ, Franklin BA, Johnson CM, et al. Physical activity and exercise recommendations for stroke survivors: A statement for healthcare professionals from the American Heart Association/American Stroke Association. Stroke. 2014;45:2532–2553.

2. Maclean N, Pound P, Wolfe C, Rudd A. The concept of patient motivation: A qualitative analysis of stroke professionals’ attitudes. Stroke. 2002;33:444–448.

3. Rapoliene J, Endzelyte E, Jaseviciene I, Savickas R. Stroke patients motivation influence on the effectiveness of occupational therapy. Rehabil Res Pract. 2018;2018:9367942.

4. McGrane N, Galvin R, Cusack T, Stokes E. Addition of motivational interventions to exercise and traditional physiotherapy: A review and meta-analysis. Physiotherapy. 2015;101:1–12.

5. Oyake K, Suzuki M, Otaka Y, Tanaka S. Motivational strategies for stroke rehabilitation: A descriptive cross-sectional study. medRxiv. 2019:19011023.

6. Dobkin BH, Plummer-D’Amato P, Elashoff R, Lee J, SIRROWS Group. International randomized clinical trial, stroke inpatient rehabilitation with reinforcement of walking speed (SIRROWS), improves outcomes. Neurorehabil Neural Repair. 2010;24:235–242.

7. Cheng D, Qu Z, Huang J, Xiao Y, Luo H, Wang J. Motivational interviewing for improving recovery after stroke. Cochrane Database Syst Rev. 2015:CD011398.

8. Forster A, Brown L, Smith J, House A, Knapp P, Wright JJ, et al. Information provision for stroke patients and their caregivers. Cochrane Database Syst Rev. 2012;11:CD001919.

9. Hohmann E, Brand JC, Rossi MJ, Lubowitz JH. Expert opinion is necessary: Delphi panel methodology facilitates a scientific approach to consensus. Arthroscopy. 2018;34:349–351.

10. Junger S, Payne SA, Brine J, Radbruch L, Brearley SG. Guidance on conducting and reporting Delphi studies (CREDES) in palliative care: Recommendations based on a methodological systematic review. Palliat Med. 2017;31:684–706.

11. Diamond IR, Grant RC, Feldman BM, Pencharz PB, Ling SC, Moore AM, et al. Defining consensus: A systematic review recommends methodologic criteria for reporting of Delphi studies. J Clin Epidemiol. 2014;67:401–409.

12. von der Gracht HA. Consensus measurement in Delphi studies: Review and implications for future quality assurance. Technol Forecast Soc Chang. 2012;79:1525–1536.

13. Hasson F, Keeney S, McKenna H. Research guidelines for the Delphi survey technique. J Adv Nurs. 2000;32:1008–1015.

14. Terwee CB, Prinsen C, Chiarotto A, de Vet H, Bouter LM, Alonso J, et al. COSMIN methodology for assessing the content validity of PROMs–user manual version 1.0. 2018. https://www.cosmin.nl/wp-content/uploads/COSMIN-methodology-for-content-validity-user-manual-v1.pdf. Accessed February 23, 2020.

15. Guilloteaux MJ, Dörnyei Z. Motivating language learners: A classroom-oriented investigation of the effects of motivational strategies on student motivation. TESOL Quarterly. 2008;42:55–77.

16. Pessiglione M, Vinckier F, Bouret S, Daunizeau J, Le Bouc R. Why not try harder? Computational approach to motivation deficits in neuro-psychiatric diseases. Brain. 2018;141:629–650.

17. Oyake K, Kondo K, Tanaka S. Categorization of motivational strategies in rehabilitation based on arcs model. Proceedings of the 34th Annual Conference of Japan Society for Educational Technology. 2018:667–668.

18. Maclean N, Pound P, Wolfe C, Rudd A. Qualitative analysis of stroke patients’ motivation for rehabilitation. BMJ. 2000;321:1051–1054.

19. Dobkin BH. Behavioral self-management strategies for practice and exercise should be included in neurologic rehabilitation trials and care. Curr Opin Neurol. 2016;29:693–699.

20. Simpson LA, Eng JJ, Tawashy AE. Exercise perceptions among people with stroke: Barriers and facilitators to participation. Int J Ther Rehabil. 2011;18:520–530.

21. Rimmer JH, Wang E, Smith D. Barriers associated with exercise and community access for individuals with stroke. J Rehabil Res Dev. 2008;45:315–322.

22. Nicholson S, Sniehotta FF, van Wijck F, Greig CA, Johnston M, McMurdo ME, et al. A systematic review of perceived barriers and motivators to physical activity after stroke. Int J Stroke. 2013;8:357–364.

23. Nicholson SL, Greig CA, Sniehotta F, Johnston M, Lewis SJ, McMurdo ME, et al. Quantitative data analysis of perceived barriers and motivators to physical activity in stroke survivors. J R Coll Physicians Edinb. 2017;47:231–236.

24. Damush TM, Plue L, Bakas T, Schmid A, Williams LS. Barriers and facilitators to exercise among stroke survivors. Rehabil Nurs. 2007;32:253-260, 262.

25. Kelley K, Clark B, Brown V, Sitzia J. Good practice in the conduct and reporting of survey research. Int J Qual Health Care. 2003;15:261–266.

26. McGrane N, Cusack T, O’Donoghue G, Stokes E. Motivational strategies for physiotherapists. Physl Ther Rev. 2014;19:136–142.

27. De Vet E, Brug J, De Nooijer J, Dijkstra A, De Vries NK. Determinants of forward stage transitions: A Delphi study. Health Educ Res. 2005;20:195–205.

28. Ahuja M, Aseltine R, Warren N, Reisine S, Williams PH, Cislo A. Challenges faced with the implementation of web-based data query systems for population health: Development of a questionnaire based on expert consensus. Pilot Feasibility Stud. 2018;4:113.

29. Atkins D, Best D, Briss PA, Eccles M, Falck-Ytter Y, Flottorp S, et al. Grading quality of evidence and strength of recommendations. BMJ. 2004;328:1490.

30. Abrams P, Khoury S. International consultation on urological diseases: Evidence-based medicine overview of the main steps for developing and grading guideline recommendations. Neurourol Urodyn. 2010;29:116–118.

31. Winstein CJ, Stein J, Arena R, Bates B, Cherney LR, Cramer SC, et al. Guidelines for adult stroke rehabilitation and recovery: A guideline for healthcare professionals from the American Heart Association/American Stroke Association. Stroke. 2016;47:e98–e169.

32. Pan L, Song A, Wang S, Duan S. Experimental study on upper-limb rehabilitation training of stroke patients based on adaptive task level: A preliminary study. Biomed Res Int. 2019;2019:2742595.

33. Sugavanam T, Mead G, Bulley C, Donaghy M, van Wijck F. The effects and experiences of goal setting in stroke rehabilitation - a systematic review. Disabil Rehabil. 2013;35:177–190.

34. Sonoda S, Saitoh E, Nagai S, Kawakita M, Kanada Y. Full-time integrated treatment program, a new system for stroke rehabilitation in Japan: Comparison with conventional rehabilitation. Am J Phys Med Rehabil. 2004;83:88–93.

35. Ryan RM, Deci EL. Intrinsic and extrinsic motivations: Classic definitions and new directions. Contem Educ Psychol. 2000;25:54–67.

36. Prakash V, Shah MA, Hariohm K. Family’s presence associated with increased physical activity in patients with acute stroke: An observational study. Braz J Phys Ther. 2016;20:306–311.

37. Michimata A, Suzukamo Y, Izumi SI. Development of clinicians’ communication skills influences the satisfaction, motivation, and quality of life of patients with stroke. Int J Phys Med Rehabil. 2013;1:174.

38. Laver KE, Lange B, George S, Deutsch JE, Saposnik G, Crotty M. Virtual reality for stroke rehabilitation. Cochrane Database Syst Rev. 2017;11:CD008349.

39. Popović MD, Kostić MD, Rodić SZ, Konstantinović LM. Feedback-mediated upper extremities exercise: Increasing patient motivation in poststroke rehabilitation. Biomed Res Int. 2014;2014:520374.

40. Morris J, Oliver T, Kroll T, Macgillivray S. The importance of psychological and social factors in influencing the uptake and maintenance of physical activity after stroke: A structured review of the empirical literature. Stroke Res Treat. 2012;2012:195249.

41. Japanese Physical Therapy Association. http://www.japanpt.or.jp/english/international/for-foreigner/english/. Accessed Fubruary 23, 2020.

42. Japanese Association of Occupational Therapists. http://www.jaot.or.jp/en/resource.html. Accessed February 23,2020.

43. Japanese Association of Speech-Language Therapists. https://www.japanslht.or.jp/english/. Accessed February 23, 2020.

44. The Japanese Association of Rehabilitation Medicine. http://www.jarm.or.jp/rjn/data/. Accessed February 23, 2020.

